# Type I, II, and III interferon signatures correspond to COVID-19 disease severity

**DOI:** 10.1101/2021.03.10.21253317

**Authors:** Myung-Ho Kim, Shadi Salloum, Jeffrey Y Wang, Lai Ping Wong, James Regan, Kristina Lefteri, Zachary Manickas-Hill, MGH COVID-19 Collection & Processing Team, Jonathan Z Li, Ruslan I Sadreyev, Xu G Yu, Raymond T Chung

## Abstract

We analyzed the plasma levels of interferons and cytokines, and the expression of interferon-stimulated genes in peripheral blood mononuclear cells in COVID-19 patients with different disease severity. Mild patients exhibited transient type I interferon responses, while ICU patients had prolonged type I interferon responses with hyper-inflammation mediated by interferon regulatory factor 1. Type II interferon responses were compromised in ICU patients. Type III interferon responses were induced in the early phase of SARS-CoV-2 infection, even in convalescent patients. These results highlight the importance of type I and III interferon responses during the early phase of infection in controlling COVID-19 progression.

## Background

Infection with the severe acute respiratory syndrome coronavirus 2 (SARS-CoV-2) results in diverse clinical outcome of coronavirus disease 2019 (COVID-19). Most COVID-19 patients experience mild clinical course, but approximately 5 % of the SARS-CoV-2 positive patients experience severe disease [1], acute respiratory distress syndrome, which necessitates supplemental oxygen therapy or intensive care unit (ICU) care. Patients with mild cases, who do not need either hospitalization or ICU care, recover within around 14 days after symptom onset with viral clearance [2]. However, severe patients who need ICU care experience mild to moderate symptoms followed by a secondary respiratory worsening with prolonged viral load. Although the vaccination against SARS-CoV-2 has been feasible, still therapeutic options for COVID-19 patients are limited: SARS-CoV-2 neutralizing antibodies, antiviral agents, immunosuppressive agents.

Interferon (IFN) responses constitute the major defense against SARS-CoV-2 infection. Virus recognition by innate immune sensors of host cell induces type I and III IFN production. Type I and Type III IFNs induce the expression of interferon-stimulated genes (ISGs) which have the antiviral capacity [3]. Type I IFN, not Type III IFN, induces proinflammatory genes’ expression by selective induction of the transcription factor interferon regulatory factor 1 (IRF-1) [4]. In contrast to virally induced Type I and III IFN, Type II IFN is produced predominantly by T and NK cells upon stimulation with antigens and cytokines. Type II IFN stimulates antigen-specific adaptive immunity and activates innate immunity, particularly through the activation of macrophages.

However, SARS-CoV-2 evades the IFN responses of host by escaping immune recognition, suppressing the functions of IFNs and ISGs, and interfering antigen presentation process [3]. IFN responses modulated by both viral and host factors determine the clinical outcome of COVID-19 patients. Several studies have reported the impaired type I and II IFN responses in severe COVID-19 patients [5, 6], still the dynamic IFN responses during SARS-CoV-2 infection needs to be defined. Here, we comprehensively investigated type I, II, and III IFN signatures in COVID-19 with different disease severity. We analyzed the plasma levels of IFNs and IRF-1 regulated cytokines/chemokines, and the expression of ISGs in peripheral blood mononuclear cells (PBMCs).

## Methods

### Subjects and specimen collection

Patients were recruited between March and June 2020 at Massachusetts General Hospital by the Massachusetts Consortium on Pathogen Readiness. Patients were diagnosed with COVID-19 by reverse transcription polymerase chain reaction (RT-PCR) testing of SARS-CoV-2 on a nasopharyngeal swab. The severity of COVID-19 was classified based on the National Institute of Health COVID-19 treatment guideline. The study conforms to the principles outlined in the Declaration of Helsinki and received approval by the Ethics Committees of the Massachusetts General Hospital, Boston, USA (approval number: 2020P000804) All participants provided informed consent.

### Plasma Cytokine and Chemokine Analysis

The plasma levels of IFN-α (41110-1, R&D Systems, Minneapolis, MN) and IFN-λ1/3 (DY1598B-05, R&D Systems) were measured by enzyme-linked immunosorbent assay. The plasma levels of IFN-γ, IL-6, IL-12 (p70), IL-18, TNF-α, TRAIL, CCL2, CCL4, CCL7, CCL8, CXCL8, CXCL9, and CXCL10 were measured using the MILLIPLEX^®^ Human Cytokine/Chemokine/Growth Factor Panel A (HCYTA-60K, Merck Millipore, Billerica, MA) and Panel II (HCYP2MAG-62K, Merck Millipore).

### Interferon-stimulated genes analysis

Interferon-stimulated genes (ISGs), previously reported to be relevant in viral infection [4, 7], were selected for analyzing mRNA expression of ISGs. mRNA expression of ISGs was quantified by the NanoString platform (NanoString Technologies, Seattle, WA) and RT-PCR using PowerUP™ SYBR™ Green Master Mix (Thermo Fisher Scientific, Waltham, MA) **(Supplementary Table 2)**.

### Statistics

Grouped data are generally presented as median ± IQR, with groups compared by the Kruskal-Wallis test with Dunn’s multiple comparisons test for non-parametric data using Prism 9.0 (Graphpad)

## Supporting information

Supplementary Figures, Tables, Acknowledgment

## Data Availability

Not applicable

## Results

### Patients and Sample collection

Patients confirmed positive for SARS-CoV-2 were subdivided into three groups based on disease severity during their clinical encounters: Outpatient (Out, n=23), Hospitalization under non-ICU conditions (Mild, n=21), Hospitalization in the ICU (ICU, n=23). Convalescent patients, who were recovered from COVID-19 and confirmed negative for SARS-CoV-2, were included as a control group (Conv, n=19). The blood samples were collected typically between 5 to 44 days after symptom onset (DfSO). **(Supplementary Table 1 and Supplementary Figure 1A)**.

### The plasma levels of type I, II, and III interferons in COVID-19 patients

The plasma levels of the type I IFN IFN-α were the highest in the ICU patients, followed by the Mild and Out patients. However, when evaluated by duration after symptom onset, some of the Out and Mild patients at less than 14 DfSO exhibited higher levels of IFN-α than ICU patients. In contrast, at 15-26 DfSO, most of the Mild and Out patients had decreased levels of IFN-α, while ICU patients still had increased levels of IFN-α **(Figure 1A)**. The plasma levels of type II IFN IFN-γ were significantly reduced in ICU patients regardless of sample collection date **(Figure 1B)**. The plasma levels of the type III IFN IFN-λ1/3 were comparable among patients. It appeared in Mild and ICU patients that the IFN-λ1/3 was preferentially induced during the early phase of infection **(Figure 1C)**.

**Figure 1.**
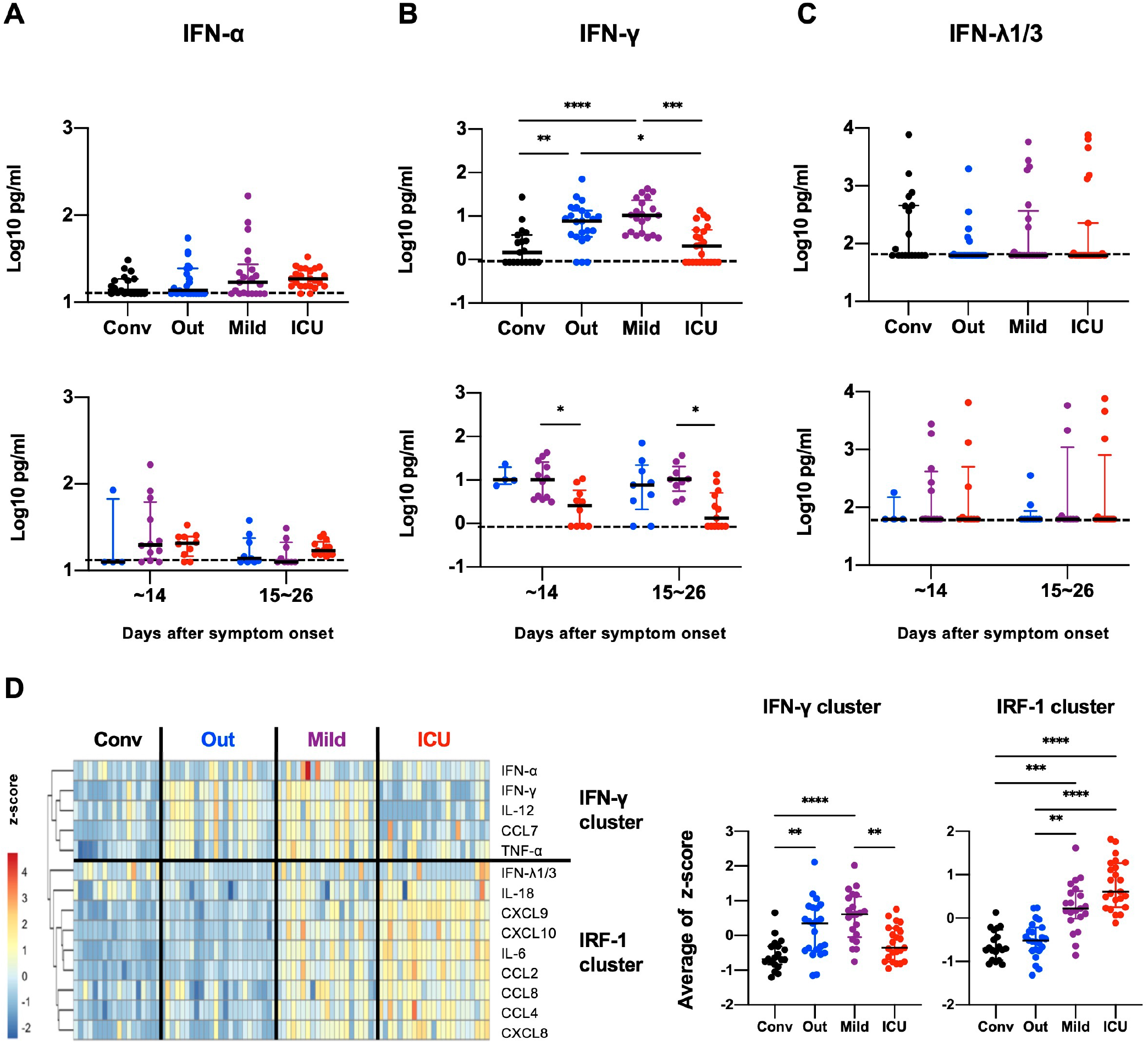
The plasma levels of type I, II, and Ill interferons in COVID-19 patients. The plasma concentration of **(A)** IFN-α, **(B)** IFN-γ, and **(C)** IFN-λ1/3 of each disease severity (Conv: Convalescence, n= 19; Out: Outpatients, n= 23; Mild: Hospitalization under non-ICU condition, n= 23; ICU: Hospitalization in ICU, n= 23). The levels of IFN-α, IFN-γ, and IFN-λ1/3 were subdivided into within 14 days after symptom onset (DfSO) and between 15 to 26 DfSO. The detection limits are indicated by dotted lines. **(D)** Heatmap clustering of the plasma levels of IRF-1 related cytokines and chemokines. The value of concentration was transformed into z-score for heatmap analysis. Heatmap clustering of plasma cytokines and chemokines yielded two major clusters: IFN-γ cluster and IRF-1 cluster. z-scores of cytokines and chemokines belonging to IFN-γ cluster or IRF-1 cluster were averaged in each patient and then compared by disease severity. Significance testing among groups was performed using Kruskal-Wallis test with Dunn’s multiple comparisons test. * P<0.05, ** P<0.01, *** P<0.001, **** P<0.0001.

### The plasma levels of IRF-1 regulated cytokines and chemokines

Levels of most of the IRF-1 regulated cytokines and chemokines, including IL-6, IL-18, TNF-α, CCL2, CCL4, CCL8, CXCL8, CXCL9, and CXCL10, were highest in the ICU patients, followed by the Mild and Out patients in a similar pattern to the IFN-α levels **(Supplementary Figure 2A)**. The levels of IL-12, CCL7, and TRAIL were higher in Out and Mild patients, as with IFN-γ **(Supplementary Figure 2B)**. Heatmap clustering of plasma cytokines and chemokines yielded two major clusters: one consisting of IFN-γ, IL-12, CCL7, and TNF-α (IFN-γ cluster) and another cluster consisting of IRF-1 regulated cytokines and chemokines (IRF-1 cluster). IFN-γ cluster was upregulated in the Out and Mild patients, but not in the ICU patients **(Figure 1D)**.

### The expression of ISGs in PBMCs

IRF-1 regulated genes in PBMCs were upregulated in only the ICU patients, while antiviral genes were upregulated in both Mild and ICU patients **(Figure 2A**,**B and Supplementary Figure 3)**. The expression of antiviral genes and IRF-1 regulated genes was positively correlated with plasma IFN-α levels but was not associated with the type II IFN or type III IFN levels **(Figure 2C and Supplementary Figure 4)**. The IFN-γ stimulated genes, including *HLA-A, HLA-B, B2M, HLA-DPA1, HLA-DRA*, and *CIITA*, were downregulated in the Mild patients but especially in the ICU patients compared to Out and Conv patients **(Figure 2D)**.

**Figure 2.**
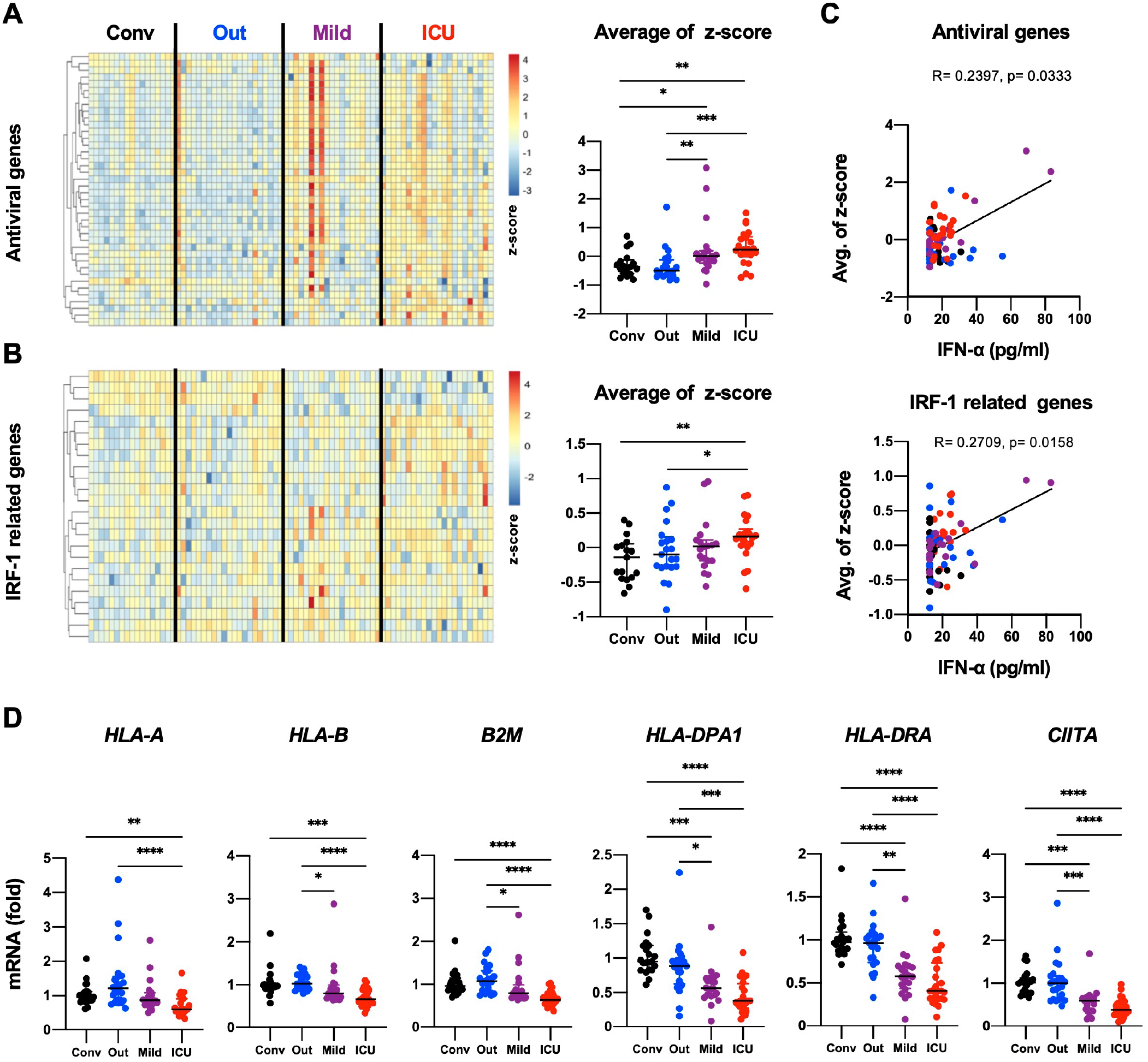
The expression of interferon-stimulated genes in PBMCs. Normalized mRNA expression levels of antiviral genes **(A)** and IRF-1 related genes **(B)** in PBMCs were transformed into z-score and visualized in the form of heatmap. Average z-scores of antiviral genes or IRF-1 related genes were compared by disease severity. **(C)** Correlation between the IFN-α levels and the average z-scores of antiviral genes or IRF-1 related genes. Spearman’s rank test was used for the correlations. The regression line (solid) are shown. **(D)** mRNA expression levels of IFN-γ stimulated genes. Significance testing among groups was performed using Kruskal-Wallis test with Dunn’s multiple comparisons test.* P<0.05, ** P<0.01, *** P<0.001, **** P<0.0001.

## Discussion

It is known that older adults and men are at higher risk of hospitalization and death if they are diagnosed with COVID-19. Our study further affirmed this finding since the Mild and ICU patients were older than Out patients **(Supplementary Figure 1B)**. However, Mild and ICU patients displayed differences in IFN signatures despite sharing comparable ages. The plasma levels of type I, II, and III IFNs were not correlated with age in the Mild and ICU patients, while IL-6 levels correlated with age **(Supplementary Figure 5)**. Similarly, levels of type I, II, and III IFNs and IL-6 within each group were not significantly different between genders and between White/Non-Hispanic and the other race/ethnicities **(Supplementary Figure 6A and B)**. Therefore, we conclude that our observations were not explained by age, gender, or race/ethnicity.

Several studies have reported that the type I and III IFN responses of severe COVID-19 patients are suppressed during the early phase of infection [5, 8]. However, other studies have shown that severe COVID-19 patients have robust type I IFN responses [6, 9]. In our study, we observed the induction of rapid and transient type I IFN responses in Out and Mild patients, but the prolonged type I IFN responses in ICU patients. Furthermore, lower viral loads and the shorter duration of hospitalization of Out and Mild patients suggest rapid virus clearance, while the ICU patients displayed prolonged hospitalization and higher viral loads **(Supplementary Figure 1C and D)**.

Type I IFN activates transcription of proinflammatory genes by inducing the transcription factor IRF-1 [3, 4, 7], which upregulates cytokines that contribute to hyper-inflammation in COVID-19. In our study, the ICU patients had higher plasma IFN-α levels during the later phase of infection. The expression of IRF-1 regulated genes in PBMCs was upregulated in only the ICU patients and positively correlated with plasma IFN-α levels. These results demonstrate that the hyper-inflammation in ICU patients can be traced to prolonged type I IFN responses.

Several studies have reported the reduction of the plasma type II IFN in severe COVID-19 patients similar to our findings [5]. A series of analyses on the immune cells have demonstrated that IFN-γ producing CD4+T, CD8+T, and NK cells are exhausted and depleted in severe COVID-19 patients [10, 11], which could plausibly explain the decreased plasma IFN-γ levels in ICU patients. To restore the exhausted immune cells, several studies are testing the use of anti PD-1 antibody camrelizumab (ChiCTR2000029806) and recombinant IL-2 (ChiCTR2000030167) against COVID-19.

The levels of IFN-γ stimulated genes were somewhat diminished in Mild patients compared with Out and Conv patients, they were still substantially upregulated compared to ICU patients. In this regard, these findings were similar to the pattern observed in plasma IFN-γ levels. Viruses including coronaviruses, MERS-CoV and H5N1 influenza virus, interfere with the antigen presentation process through MHC molecules. The ORF8 protein of SARS-CoV-2 downregulates MHC class I molecules [12], although the evidence for the interference of antigen presentation by SARS-CoV-2 is still lacking. Several studies have demonstrated the downregulation of MHC class I and II molecules in antigen presenting cells of COVID-19 patients, regardless of disease severity [13, 14]. Thus, decreased expression of MHC molecules in PBMCs and reduced plasma IFN-γ could synergistically subvert adaptive immunity in ICU patients.

Type III IFN is induced earlier than type I IFN upon virus infection, and suppresses initial viral spread without activating inflammation. Type I IFN response is triggered later to enhance antiviral activity and induce IRF-1 mediated inflammatory responses [3, 7]. Interestingly, some of the Conv patients showed increased IFN-λ1/3 levels **(Figure 1C)** and the upregulated antiviral genes, while not inducing IRF-1 related genes **(Figure 2A and B)**, even though the patients were confirmed negative for SARS-CoV-2. While it is remotely possible that these patients may have been re-infected with SARS-CoV-2 and not developed detectable viral RNA, it would appear much more likely that the virus was rapidly cleared by type III IFN responses prior to engagement of a type I IFN responses.

Type III IFN therapy could be a novel therapeutic strategy against COVID-19. Early use of type I IFNs has benefits in virus clearance and clinical outcomes in COVID-19 patients. However, later use of type I IFNs could potentially delay recovery and increase mortality [15]. This could be attributed to IRF-1 related hyper-inflammation. While the expression of type I IFN receptors is ubiquitous, the expression of type III IFN receptors is limited to epithelial cells [3]. Thus, type III IFN therapy could be an effective alternative to type I IFN therapy since it can promote virus clearance without inducing IRF-1 related inflammation. Several clinical trials by our group and others are ongoing to confirm the validity of IFN-λ therapy against COVID-19 in both mild and severe patients (NCT04343976, NCT04354259, NCT04388709, and NCT04344600). Two independent clinical trials testing the IFN-λ therapy in outpatients with COVID-19 collectively indicate an antiviral effect in ambulatory patients with COVID-19 with high levels of virus.

Overall, our analyses provide a much clearer picture of the dynamic signatures of type I, II, and III IFN during COVID-19. Type I and III IFN responses during the early phase of infection appear to be important in controlling viral spread and disease progression. Failure to limit viral spread during the early phase of infection can lead to susceptibility to hyper-inflammation mediated by prolonged type I IFN and IRF-1 mediated responses, the exhaustion and depletion of IFN-γ producing cells, and the downregulation of antigen presentation through MHC class I and II. Analogously, enhanced antigen presentation and type II IFN responses appear to be associated with milder clinical illness. Finally, these findings provide rationale for use of type III IFN therapy to maximize antiviral activity without IRF-1 mediated proinflammatory responses relatively early in COVID-19 illness.

