## Supplementary Figures, Tables, Acknowledgment for "Type I, II, and III interferon signatures correspond to COVID-19 disease severity"

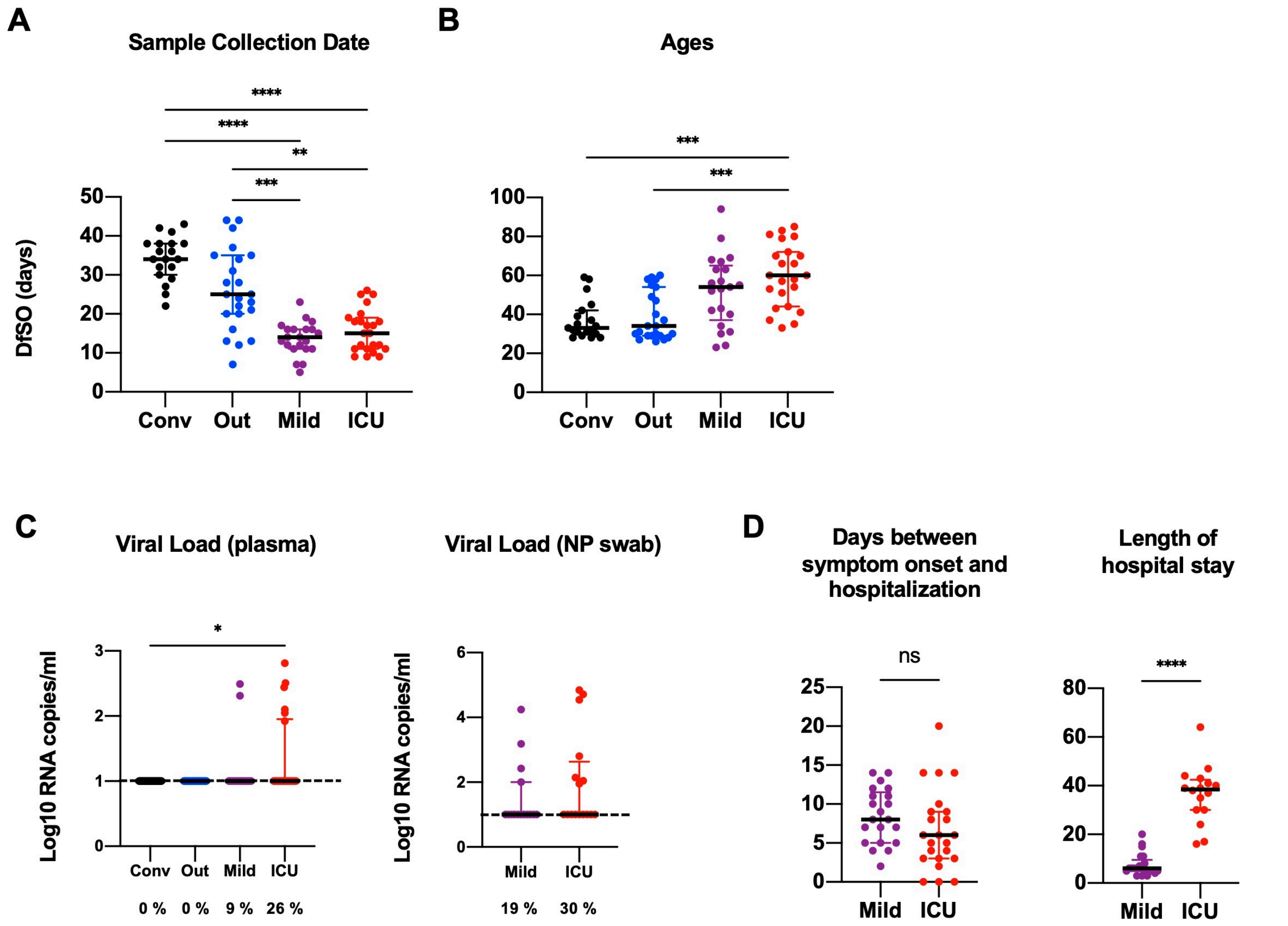


**Supplementary Figure 1. Characteristics of patients**

**(A)** Sample collection date is defined as days after symptom onset (DfSO) **(B)** Ages of patients. **(C)** SARS-CoV-2 Viral loads from plasma and nasopharyngeal swab (NP swab) of patients. The levels of viral load in plasma and nasopharyngeal swab were evaluated by quantitative reverse transcription polymerase chain reaction. The detection limits are indicated by dotted lines. The percent of samples with detectable viral loads are shown at the bottom. **(D)** Days between symptom onset and hospitalization, and length (days) of hospital stay of Mild and ICU patients. Significance testing among groups was performed using Kruskal-Wallis test with Dunn's multiple comparisons test.* P<0.05, ** P<0.01, *** P<0.001, **** P<0.0001.

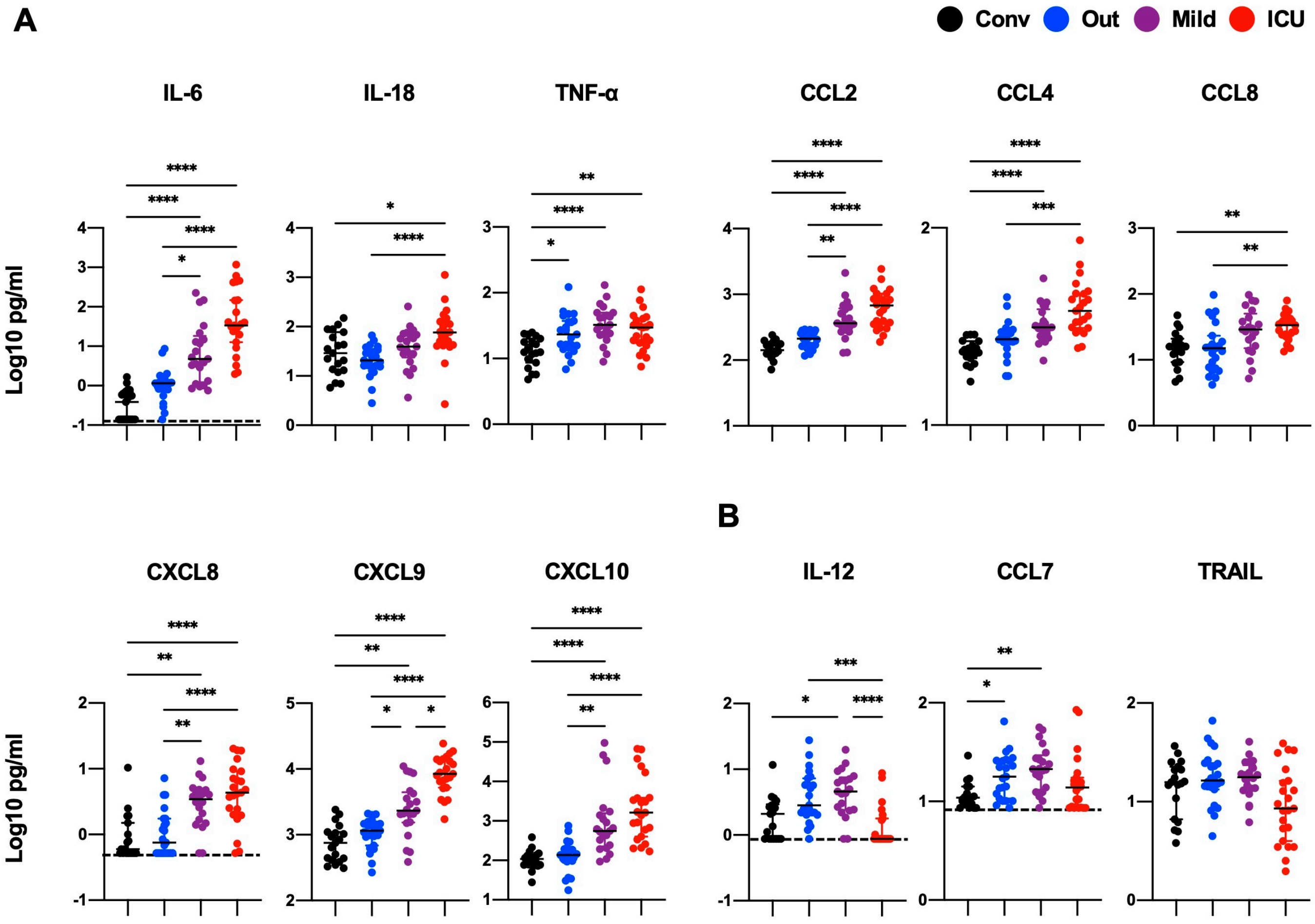


**Supplementary Figure 2. Plasma levels of interferon regulatory factor 1 (IRF-1)-regulated cytokines and chemokines.**

The detection limits are indicated by dotted lines. Significance testing among groups was performed using Kruskal-Wallis test with Dunn's multiple comparisons test. * P<0.05, ** P<0.01, *** P<0.001, **** P<0.0001.

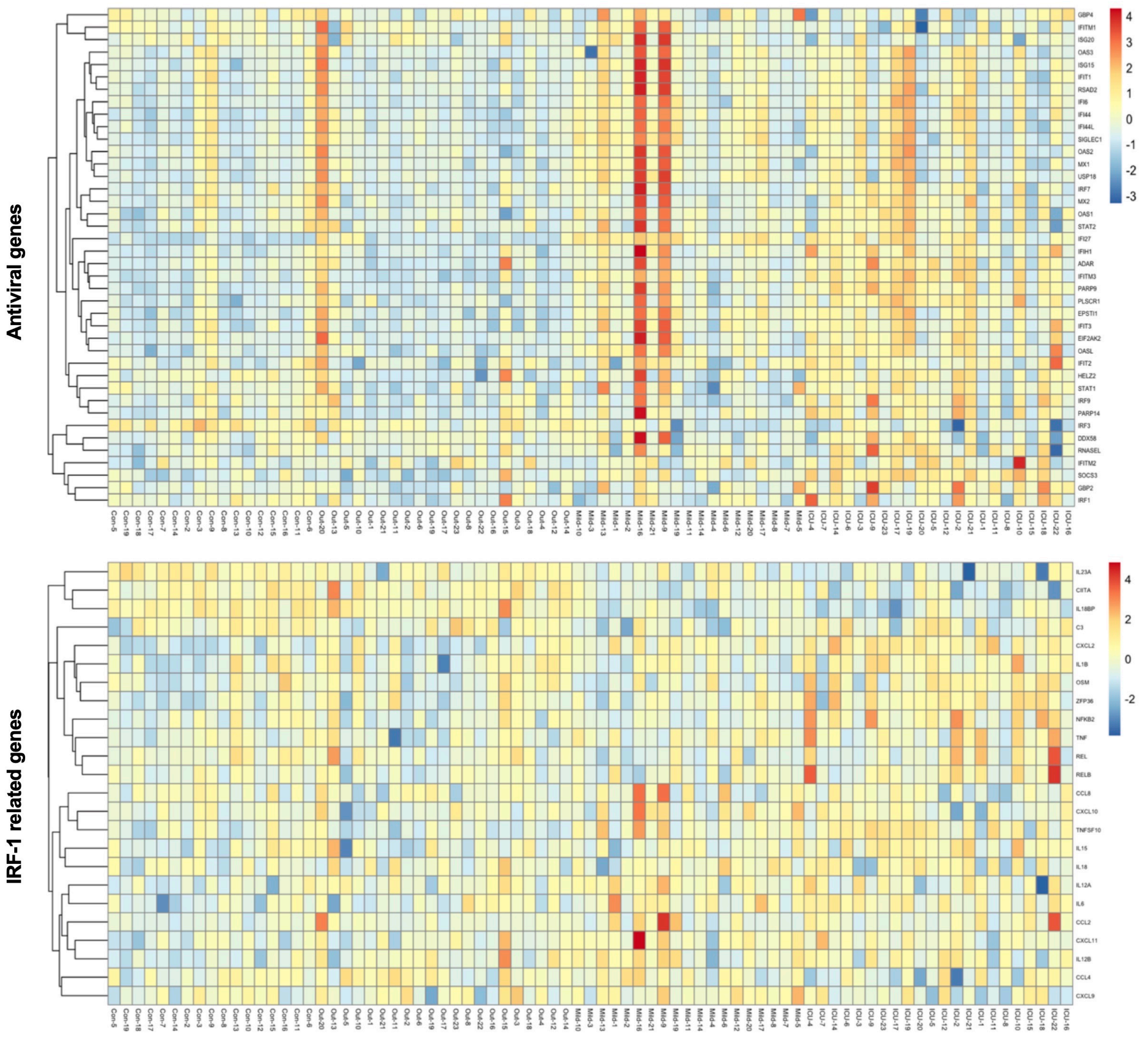


**Supplementary Figure 3. The expression of interferon-stimulated genes in PBMCs**
Normalized mRNA expression levels of antiviral genes and IRF-1 related genes in PBMCs were transformed into z-score and visualized in the form of heatmap.

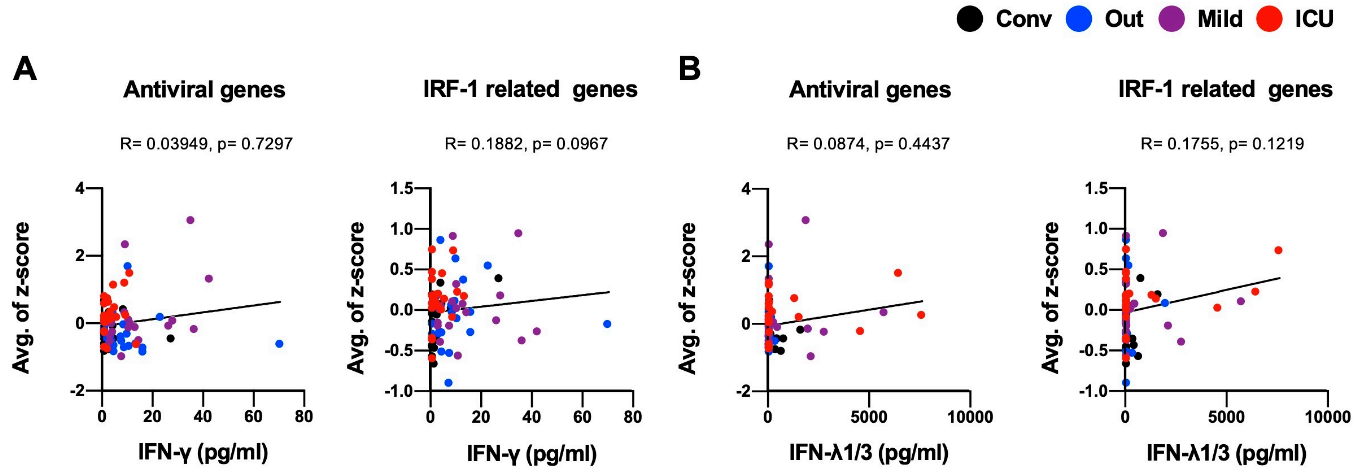


**Supplementary Figure 4. Correlation between the interferons levels and ISGs expression.**
Correlation between the levels of IFN-α **(A)**, IFN-λ1/3 **(B)** and the average z-scores of antiviral genes and IRF-1 related genes. Spearman's rank test was used for the correlations. The regression line (solid) are shown.

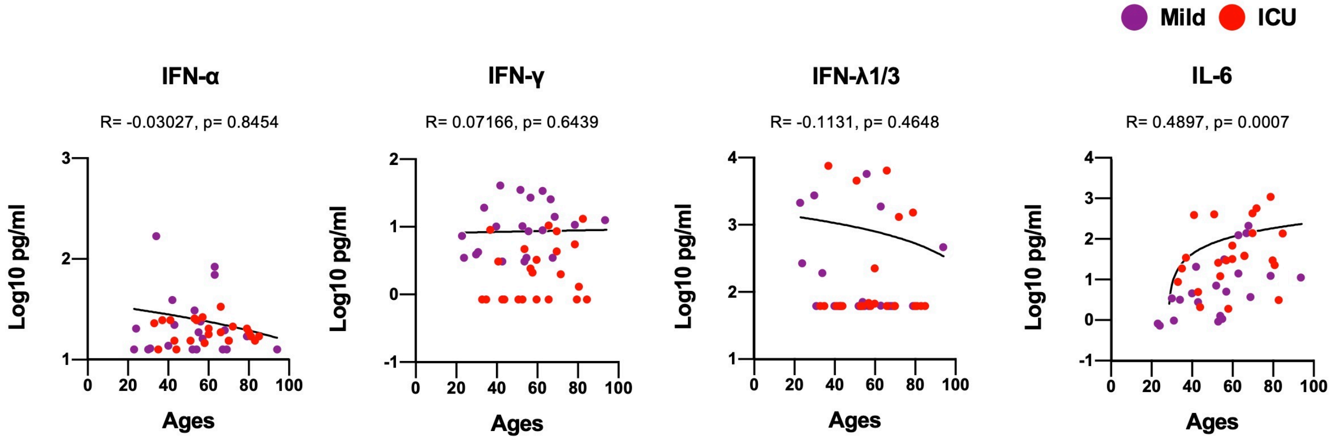


**Supplementary Figure 5. Correlation between ages and the levels of interferons and IL-6.**
Correlation between ages and the levels of IFN-α, IFN-γ, IFN-λ1/3, and IL-6. Spearman's rank test was used for the correlations. The regression line (solid) are shown.

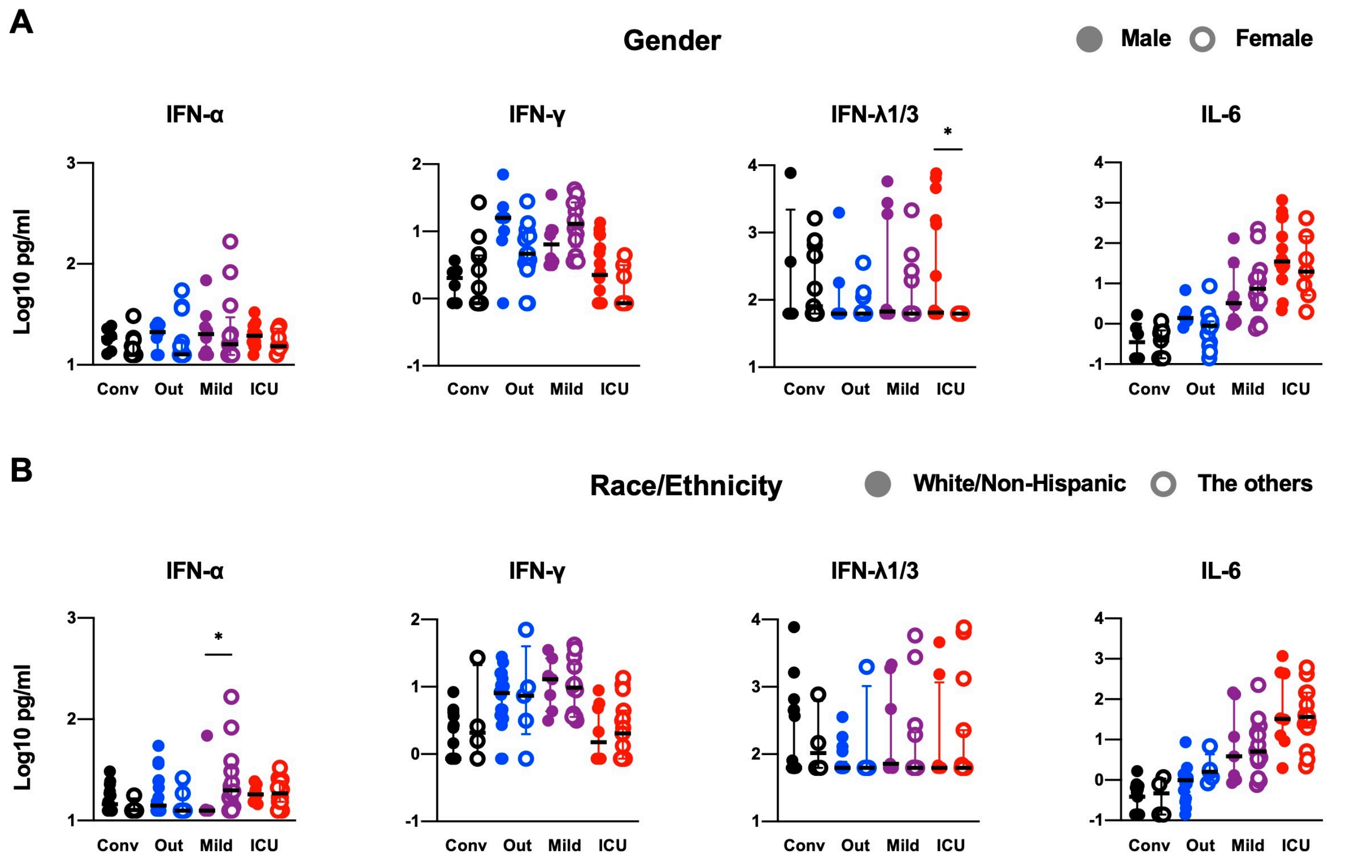


**Supplementary Figure 6. The levels of the interferons and IL-6 segmented by gender and race/ethnicity.** The levels of IFN-α, IFN-γ, IFN-λ1/3, and IL-6 were segmented by gender **(A)** and race/ethnicity **(B)**.

| Supplementary Table 1. Characteristics of patients | | | | |
|---|---|---|---|---|
| | Convalescent | Outpatients | Hospitalized | Hospitalized |
| | | | non-ICU | ICU |
| **Numbers** | 22.1% (19/86) | 26.7% (23/86) | 24.4% (21/86) | 26.7% (23/86) |
| **Ages, median (IQR)** | 33 (30-42) | 34 (29-54) | 54 (37-65) | 60 (44-72) |
| **Male, no (%)** | 6 (31.6%) | 7 (30.4%) | 8 (38.1%) | 16 (69.5%) |
| **Race/Ethnicity** | | | | |
| Asian/Non-Hispanic | 5.3 % (1/19) | 8.7 % (2/23) | - | - |
| Asian/Hispanic | - | - | 4.8 % (1/21) | - |
| Asian/Unknown | - | - | - | - |
| Black or African American/Non-Hispanic | 5.3 % (1/19) | 8.7 % (2/23) | 4.8 % (1/21) | 13.0 % (3/23) |
| Black or African American/Hispanic | - | - | 4.8 % (1/21) | 4.3 % (1/23) |
| Black or African American/Unknown | - | - | - | 4.3 % (1/23) |
| White/Non-Hispanic | 78.9 % (15/19) | 78.2 % (18/23) | 33.3 % (7/21) | 34.8 % (8/23) |
| White/Hispanic | 10.5 % (2/19) | 4.3 % (1/23) | 9.5 % (2/21) | - |
| White/Unknown | - | - | - | 4.3 % (1/23) |
| Unknown/Non-Hispanic | - | - | - | 4.3 % (1/23) |
| Unknown/Hispanic | - | - | 33.3 % (7/21) | 21.7 % (5/23) |
| Unknown/Unknown | - | - | 9.5 % (2/21) | 13.0 % (3/23) |
| **Median interval from symptom onset to admission, days (IQR)** | - | - | 8 (5-11.5) | 6 (3-9) |
| **Median duration of hospitalized, days (IQR)** | - | - | 6 (5-9.5) | 38.5 (30-42.5)* |
| **Sample collection date, days after symptom onset (IQR)** | 34 (30-38) | 25 (20-35) | 14 (11-16) | 15 (11-19) |

*the patients deceased (n=6) and currently hospitalized (n=2) at the time of analysis were excluded

**Supplementary Table 2. Primers for IFN-γ stimulated genes**

| Primer Name | Sequence (5' - >3') |
|---|---|
| *HLA-A* forward | AAA AGG AGG GAG TTA CAC TCA GG |
| *HLA-A* reverse | GCT GTG AGG GAC ACA TCA GAG |
| *HLA-B* forward | CTA CCC TGC GGA GAT CA |
| *HLA-B* reverse | ACA GCC AGG CCA GCA ACA |
| *B2M* forward | TGC TGT CTC CAT GTT TGA TGT ATCT |
| *B2M* reverse | TCT CTG CTC CCC ACC TCT AAG T |
| *HLA-DPA1* forward | ATG CGC CCT GAA GAC AGA ATG |
| *HLA-DPA1* reverse | ACA CAT GGT CCG CCT TGA TG |
| *HLA-DRA* forward | GCC AAC CTG GAA ATC ATG ACA |
| *HLA-DRA* reverse | AGG GCT GTT CGT GAG CAC A |
| *CIITA* forward | TTA TGC CAA TAT CGC GGA ACT G |
| *CIITA* reverse | CAT CTG GTC CTA TGT GCT TGA AA |

**Supplemental acknowledgments**

**MGH COVID-19 Collection & Processing Team Participants**

**Collection Team:** Kendall Lavin-Parsons[1], Blair Parry[1], Brendan Lilley[1], Carl Lodenstein[1], Brenna McKaig[1], Nicole Charland[1], Hargun Khanna[1], Justin Margolin[1], Edward DeMers[6], Kelly Judge[6], Bruce D. Walker[6], Peggy Lai[6], Musie S. Ghebremichael[6]

**Processing Team:** Anna Gonye[2], Irena Gushterova[2], Tom Lasalle[2], Nihaarika Sharma[2], Brian C. Russo[3], Maricarmen Rojas-Lopez[3], Moshe Sade-Feldman[4], Kasidet Manakongtreecheep[4], Jessica Tantivit[4], Molly Fisher Thomas[4]

**Massachusetts Consortium on Pathogen Readiness:** Betelihem A. Abayneh[5], Patrick Allen[5], Diane Antille[5], Katrina Armstrong[5], Siobhan Boyce[5], Joan Braley[5], Karen Branch[5], Katherine Broderick[5], Julia Carney[5], Andrew Chan[5], Susan Davidson[5], Michael Dougan[5], David Drew[5], Ashley Elliman[5], Keith Flaherty[5], Jeanne Flannery[5], Pamela Forde[5], Elise Gettings[5], Amanda Griffin[5], Sheila Grimmel[5], Kathleen Grinke[5], Kathryn Hall[5], Meg Healy[5], Deborah Henault[5], Grace Holland[5], Chantal Kayitesi[5], Vlasta LaValle[5], Yuting Lu[5], Sarah Luthern[5], Jordan Marchewka (Schneider)[5], Brittani Martino[5], Roseann McNamara[5], Christian Nambu[5], Susan Nelson[5], Marjorie Noone[5], Christine Ommerborn[5], Lois Chris Pacheco[5], Nicole Phan[5], Falisha A. Porto[5], Edward Ryan[5], Kathleen Selleck[5], Sue Slaughenhaupt[5], Kimberly Smith Sheppard[5], Elizabeth Suschana[5], Vivine Wilson[5], Galit Alter[6], Alejandro Balazs[6], Julia Bals[6], Max Barbash[6], Yannic Bartsch[6], Julie Boucau[6], Josh Chevalier[6], Fatema Chowdhury[6], Kevin Einkauf[6], Jon Fallon[6], Liz Fedirko[6], Kelsey Finn[6], Pilar Garcia-Broncano[6], Ciputra Hartana[6], Chenyang Jiang[6], Paulina Kaplonek[6], Marshall Karpell[6], Evan C. Lam[6], Kristina Lefteri[6], Xiaodong Lian[6], Mathias Lichterfeld[6], Daniel Lingwood[6], Hang Liu[6], Jinqing Liu[6], Natasha Ly[6], Ashlin Michell[6], Ilan Millstrom[6], Noah Miranda[6], Claire O'Callaghan[6], Matthew Osborn[6], Shiv Pillai[6], Yelizaveta Rassadkina[6], Alexandra Reissis[6], Francis Ruzicka[6], Kyra Seiger[6], Libera Sessa[6], Christianne Sharr[6], Sally Shin[6], Nishant Singh[6], Weiwei Sun[6], Xiaoming Sun[6], Hannah Ticheli[6], Alicja Trocha-Piechocka[6], Daniel Worrall[6], Alex Zhu[6], George Daley[7], David Golan[7], Howard Heller[7], Arlene Sharpe[7], Nikolaus Jilg[8], Alex Rosenthal[8], Colline Wong[8]

[1]Department of Emergency Medicine, Massachusetts General Hospital, Boston, MA, USA. [2]Massachusetts General Hospital Cancer Center, Boston, MA, USA. [3]Division of Infectious Diseases, Department of Medicine, Massachusetts General Hospital, Boston, MA, USA. [4]Massachusetts General Hospital Center for Immunology and Inflammatory Diseases, Boston, MA, USA. [5]Massachusetts General Hospital, Boston, MA, USA. [6]Ragon Institute of MGH, MIT and Harvard, Cambridge, MA, USA. [7]Harvard Medical School, Boston, MA, USA. [8]Brigham and Women's Hospital, Boston, MA, USA.
